# Statistical pitfalls of multiple exposures in causal observational studies and tools to address them

**DOI:** 10.1101/2024.02.26.24303405

**Authors:** Kevin J McIntyre, Joshua C Wiener, Emma Davies Smith

## Abstract

The Table 2 Fallacy is an interpretation error commonly encountered in medical literature. This fallacy occurs when coefficient estimates in multivariable regression models, apart from that of the primary exposure, are interpreted as total effects on the outcome. Causal diagrams can be used to identify sets of covariates that, when adjusted for, allow for unbiased estimation and correct interpretation of multiple total effects of interest. However, proper investigation of multiple total effects requires fitting several regression models and conducting multiple inferences. As the number of inferences increases, so does the rate of a false positive finding, a phenomenon known as multiplicity. While multiple comparison procedures are recognized as a critical consideration of randomized controlled trials, opinion remains divided on their use within observational studies. This commentary highlights how multiplicity may arise alongside the Table 2 Fallacy, and how causal diagrams can be used in conjunction with multiple comparison procedures to simultaneously avoid this fallacy, control the risk of spurious findings, and further align the best practices of experimental and observational studies.

## The Foundation

Randomization inhibits associations between interventions and confounders, which allows for a more accurate estimation of causal effects, thereby making randomized controlled trials (RCTs) the ‘gold standard’ study design. However, due to feasibility issues or ethical concerns, many research questions cannot be answered using RCTs. There is also an increasing wealth of electronic health data that provides a means of answering important health questions. In these scenarios, observational study designs are often conducted.

Hernán et al. (2008) proposed that observational study designs should be guided by conceptualizing an ideal, hypothetical RCT – or “target trial” – that could address the research question if it were feasible.^1^ In addition to unifying RCTs and observational studies in terms of design, the target trial framework also adopts features of RCT analysis, such as the identification of intention-to-treat and per-protocol causal contrasts. Adoption of the target trial framework has led to greater compatibility between historically conflicting results of RCTs and observational studies.^2^

Yet, opinions remain divided on some aspects of design and analysis between these two study paradigms. The control of false positive or type I error rates is increasingly recognized as critical to RCTs with multiple treatment arms, outcomes, or subgroups.^3^ On the contrary, some epidemiologists have argued that there is no or little need to control this error in observational studies,^4^ although others disagree.^5^Unlike RCTs, observational studies must employ design and analysis strategies to account for confounding. Multivariable regression can provide unbiased estimates of the total effect of a primary exposure on an outcome by adjusting for the influence of all known confounders. When the exposure of interest changes, so do the confounding pathways, and thus secondary coefficient estimates within the same model are not interpretable as total effects. Interpretation of secondary effect estimates as valid total effects was dubbed the “Table 2 Fallacy” by Westreich & Greenland (2013),^6^ and this phenomenon is common in the medical literature.^7^ This interpretation error can be avoided by ensuring that a new model is specified for each exposure of interest according to its distinct biasing pathways.

In this commentary, we highlight how multiplicity can arise alongside the Table 2 Fallacy when investigating multiple complex, causal relationships with observational data. Casual diagrams are presented as a means of avoiding the Table 2 Fallacy, and multiple comparison procedures are discussed as a means of avoiding spurious associations.

## The Tools

### Causal Diagrams

Causal diagrams, namely directed acyclic graphs (DAGs), can assist with specifying models for multiple exposures. DAGs provide investigators with a framework to explicitly state assumptions about hypothesized causal mechanisms. Within a DAG, variables are represented as nodes within a graph, and causal paths as arcs between nodes. Investigators can use DAGs to visualize confounders, and thus select covariates to include within their regression models. Lipsky and Greenland (2022) provide an accessible tutorial on how to use DAGs in medical research.^8^ Unfortunately, while DAGs are broadly accepted within the epidemiological community, they are underutilized in medical research.^9^

### Multiple Comparison Procedures

The type I error or false positive rate, denoted *α*, is the risk of falsely concluding that an association is statistically significant. The overall type I error rate is inflated when inference is performed for multiple exposures. For example, if separate tests are performed for five exposures at *α* = 0.05, the overall probability of falsely rejecting at least one hypothesis is 1 − (1 − 0.05)^5^ = 23%. As the number of tests grows, the probability of at least one false positive approaches 100%; this phenomenon is referred to as multiplicity.

Multiple comparison procedures (MCPs) adjust the significance level for each inference to prevent inflation of the overall type I error rate. For example, the Bonferroni adjustment assigns *α/K* rather than *α* to each of *K* associations to ensure that the overall type I error rate is no greater than *α*. In addition to using DAGs to properly identify models for multiple exposures, the use of MCPs can assist in minimizing the risk of spurious findings and increase the likelihood of reproducibility. However, we acknowledge that multiplicity adjustment may not always be necessary in observational studies, such as when analysis is strictly descriptive.^4^

### Statistical Significance and Interval Estimation

Over the past several decades, p-values specifically and statistical significance more broadly have received criticism for oversimplifying effect interpretation.^10^ Trial reporting guidelines like CONSORT now recommend confidence intervals be reported alongside point estimates and p-values to promote the assessment of clinical relevance in addition to statistical significance.^11^

Just as significance levels may be adjusted for multiple tests, confidence levels for multiple intervals may be adjusted to ensure simultaneous coverage. Borrowing conceptually from the Bonferroni adjustment, (1- *α*/*K*) x 100% confidence intervals can be constructed for *K* effects. These intervals provide the same information on statistical significance as *K* tests at the adjusted *α*/*K* significance level, i.e., by assessing inclusion of the null effect, but provide additional insight into effect magnitudes and directions compatible with the data. For more complex MCPs, however, the form of corresponding confidence intervals is not always straightforward. Further guidance on this topic can be found in Vickerstaff, Omar & Ambler (2019).^12^

## The Example

In Figure 1A, we present the DAG for a hypothetical causal mechanism between eight variables; dagitty^13^ and ggdagR^14^ were used for visualization and analysis, respectively. The DAG features a primary exposure (E), an outcome (O), and six covariates that are causally related to the exposure, outcome, and/or each other (A, B, C, D, F, G). There are three different combinations of covariates that, when adjusted for, produce an unbiased estimate of the total effect of E on O. These “sufficient sets” of covariates are illustrated in Figure 1B: (1) A, C, G; (2) C, D, G; and (3) F, G.

**Figure 1.**
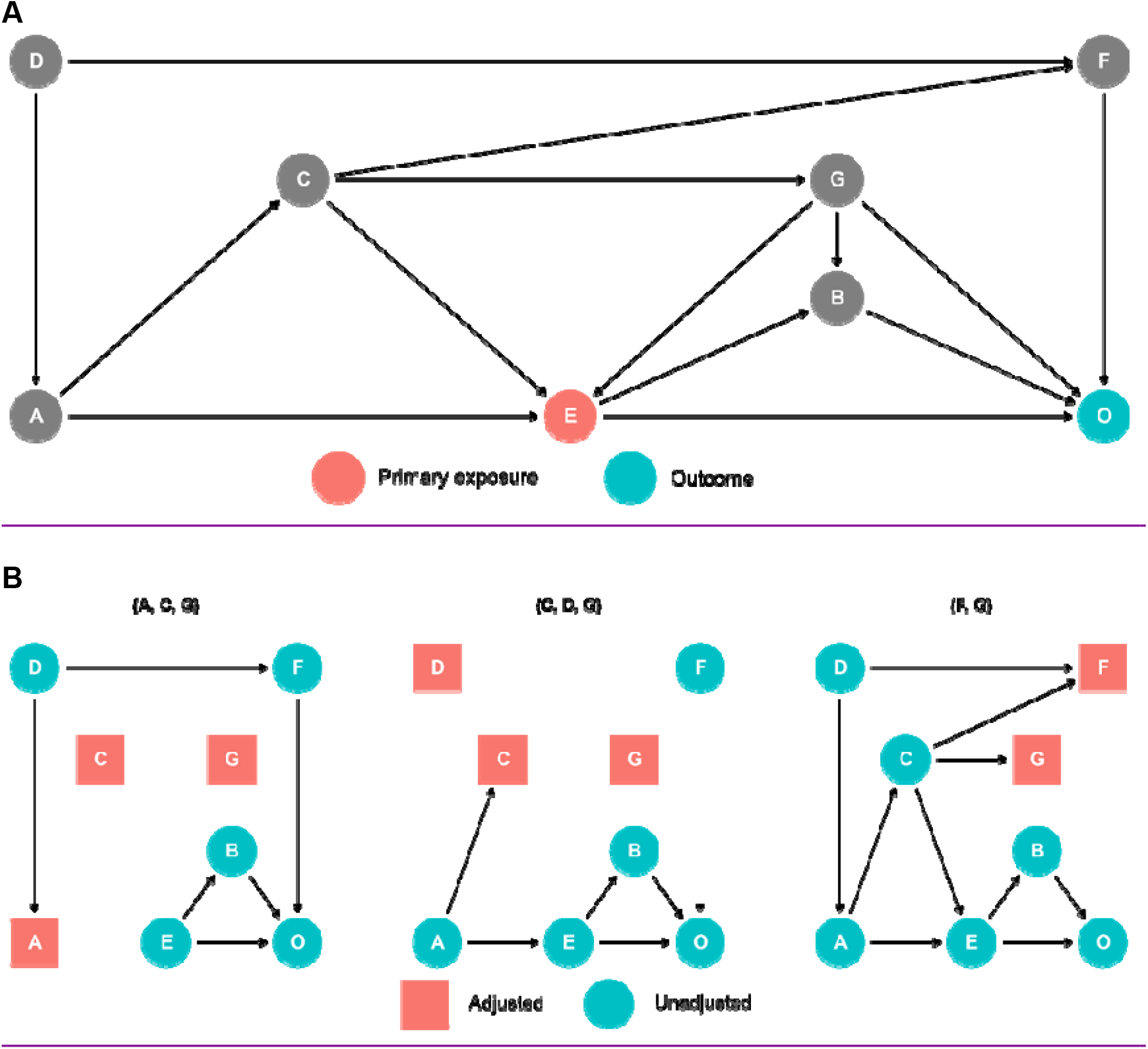
Panel A: Directed acyclic graph of hypothetical causal mechanism. Panel B: Sufficient sets for unbiased total effect of exposure E on outcome O.

Suppose an investigator fits an initial regression model conditioning on the first sufficient set (A, C, G), yielding an unbiased estimate of the total effect of E on O. If the investigator was also interested in estimating the total effect of C on O, interpreting the estimated coefficient for C within the initial model would be erroneous. The research question has changed: C is now the primary exposure, and thus the covariates needed to achieve an implied sufficient set have also changed (Figure 2A).

**Figure 2.**
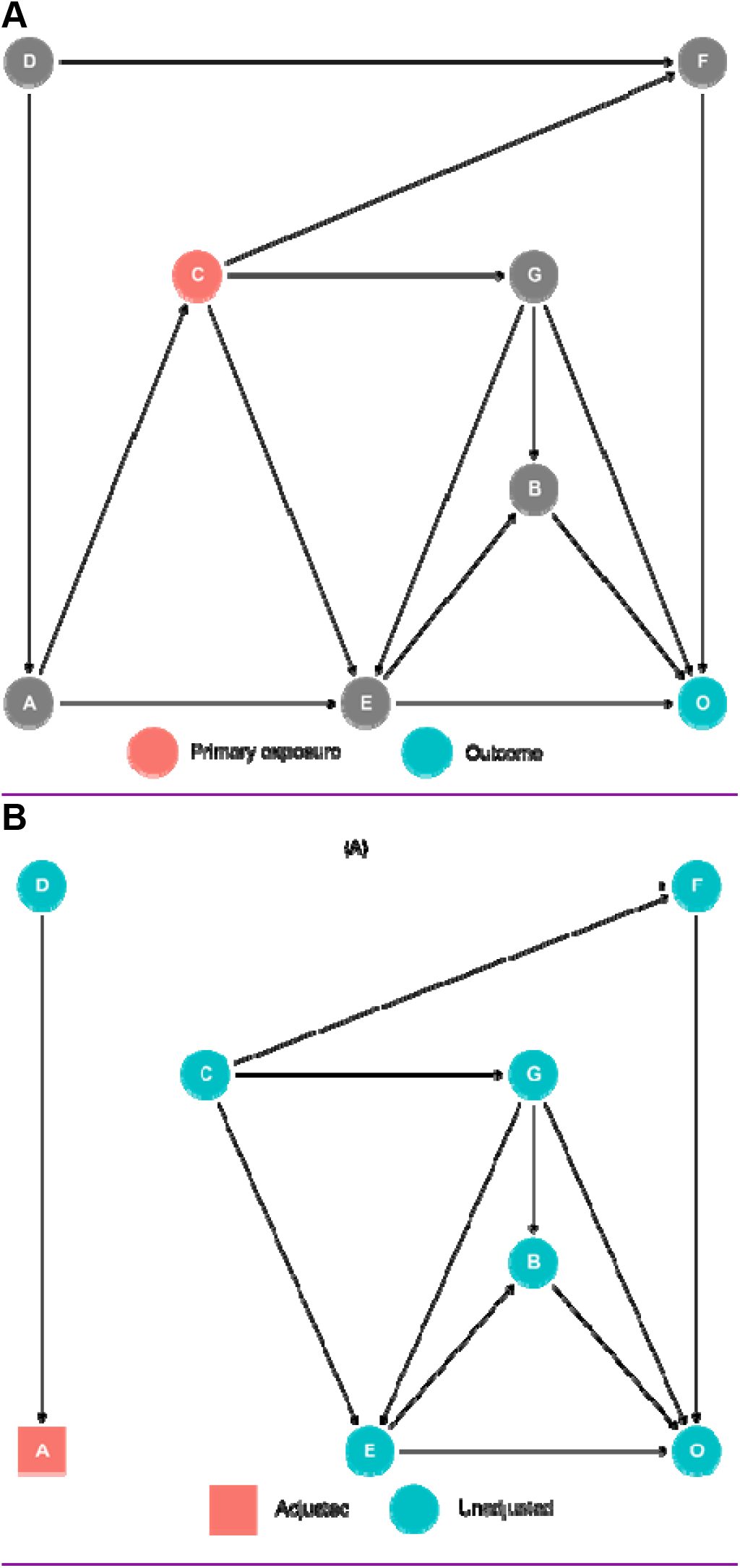
Panel A: Directed acyclic graph of hypothetical causal mechanism, with C as primary exposure. Panel B: Minimally sufficient set for unbiased total effect of C on O.

To estimate the total unbiased effect of C on O, it would only be necessary to adjust for A (Figure 2B). The effect estimate of C on O presented within a hypothetical Table 2 for E on O would not estimate the total effect, as the mediated effects of C on O through G and E are “blocked.” As such, the effect estimate of C in the initial model is guaranteed to be a biased, or rather invalid, estimate of the total effect of C on O. Similarly, if the total effect of A on O were also of interest, only adjustment for D would yield an unbiased estimate.

In this short example, we have demonstrated how three exposures can each have distinct confounding pathways, requiring separate models. If significance testing is performed for each exposure at the *α* = 0.05 level, the probability of at least one spurious finding would be approximately 14%. Using the simplest MCP, the Bonferroni correction, would ensure the overall error rate is no greater than the desired 0.05 by instead performing tests at the *α*/3 = 0.0167 level.

## The Finale

Investigation of multiple exposures is often necessary to form a complete picture of the “causal pie,” and investigators should accordingly be aware of potential pitfalls that arise in analysis. In this commentary, we highlighted two commonly overlooked statistical issues that can arise when assessing medical research questions using observational data: the Table 2 Fallacy and multiplicity. The Table 2 Fallacy occurs when coefficient estimates within a regression model, other than that of the primary exposure, are incorrectly interpreted as total effects. We demonstrated how this fallacy can be avoided by using DAGs to explicitly specify causal pathways and construct multiple models for each exposure of interest. Multiplicity, or inflation of the type I error rate, occurs when formal inference is performed for multiple associations of interest. We demonstrated how this error can be corrected using MCPs such as the Bonferroni correction. We acknowledge that MCPs are not commonly used in observational studies, despite growing adoption of RCT best practices via the target trial framework. Nonetheless, we believe that by informing practitioners of the potential for multiplicity and strategies for correction, the rigor and reproducibility of study findings can be increased, and causality-focused observational studies may better align with RCTs.

## Data Availability

All data produced in the present study are available upon reasonable request to the authors

## DECLARATIONS

## Acknowledgements

Thank you to Stephanie Armbruster, Georg Hahn, and Emma Crenshaw for their thoughtful comments which improved the quality of this manuscript (statistically significant).

## Competing Interests

The authors declare that no conflicts of interest exist with respect to the research, authorship, and/or publication of this article.

## Financial Support

The authors declare that no funds, grants, and/or other support were received during the preparation of this manuscript.

## Notes

### Competing Interest Statement

The authors have declared no competing interest.

### Funding Statement

This study did not receive any funding

### Summary of Updates

This version of the manuscript has been revised to clarify the main points addressed in the article.

